# Frequent Quantitation of Circulating Tumor Cells Predictive of Real-Time Therapy Response

**DOI:** 10.1101/2022.01.03.22268688

**Authors:** Christine M. Lim, Junli Shi, Jess Vo, Wai Min Phyo, Min Hu, Min Chin Tan, Augustine Tee, Yoon Sim Yap, Wenlong Nei, Daniel Chan, Seng Weng Wong, Meusia Neo, Norhidayah Binte Mohammad Mazian, Jackie Y. Ying, Min-Han Tan, Kaicheng Liang, Jamie Mong

## Abstract

Precision medicine is playing an increasingly important role in cancer management and treatment. Specifically in the field of oncology, circulating tumor cells (CTCs) hold significant promise in enabling non-invasive prognostication and near real-time monitoring to individualize treatments. In this study, we present strong associations between CTC subtype counts with treatment response and tumor staging in lung, nasopharyngeal and breast cancers. Longitudinal analysis of CTC count changes over short-time windows further reveals the ability to predict treatment response close to real-time. Our findings demonstrate the suitability of CTCs as a definitive blood-based metric for continuous treatment monitoring. Robust processing of high-throughput image data, explainable classification of CTC subtypes and accurate quantification were achieved using an in-house image analysis system ‘CTC-Quant’, which showed excellent agreement with expert opinion upon extensive validation.

## Introduction

The rise in precision medicine over the past decade saw a shift in cancer management from a reactive to a proactive discipline (1). Proactive cancer screening efforts worldwide (especially for high-risk individuals) have resulted in early intervention and overall decreases in cancer-specific deaths. According to the American Cancer Society (ACS) (2), age-adjusted breast cancer mortality rates have declined 40% from 1989 to 2016, with an estimated 348,800 expected deaths averted in US women; the introduction of the Papanicolaou (Pap) test in the mid-20th century resulted in a continuous decline of cervical cancer mortality rates ever since; lung cancer screening has seen mortality rates decline by 45% (men) and 19% (women) since 2002. Post-diagnosis, predictive biomarkers such as BRCA1 in Breast and ovarian cancers (3,4), KRAS in colorectal cancer (5), and PDL1 mutations in lung cancer (6) provide solid foundation for evidence-based individualized medicine seeking to deliver the most suitable treatments to each patient. Despite this, cancer metastasis is still the predominant cause of the majority of cancer deaths (7,8). Similar to screening for early intervention, early indications of disease progression and treatment resistance could potentially shape clinical decision making for each individual to increase treatment efficacy and reducing toxicity. Indeed, accurate prognosis and refined treatment monitoring could improve treatment outcomes. In this respect, circulating tumor cells (CTCs) have gained considerable attention as they are believed to be key drivers of metastatic dissemination (7,8).

The promise of CTCs as prognostic biomarkers arose due to their notable advantage of non-invasiveness and demonstrated ability to provide prognostic information. Small amounts of blood samples can be obtained routinely during clinical visits for study. Analysis of such samples have reported CTC detection and quantification to correlate with clinico-pathological parameters (9-12), associate with lower overall and relapse-free survival rates (13) and have prognostic significance in predicting treatment response or resistance (12,14,15). In addition, phenotypic subtyping of heterogeneous CTCs into epithelial CTCs (epCTCs), biphenotypic CTCs (bipCTCs), mesenchymal CTCs (mCTCs) (16) and clusters (11,17) further enhances the ability to enable more targeted analysis based on their heightened metastatic potential and association with poorer prognosis.

In this retrospective study, CTC image data (CTCs captured using microsieve) of lung, nasopharyngeal and breast cancer patient samples were analyzed. Strong associations between CTC subtype counts with treatment response and tumor staging were found. In addition, longitudinal patterns of CTC count changes over short-time windows are also reported to further enhance prediction of treatment response.

To perform objective and discriminative high-throughput analysis of CTC image data, we have developed a customized CTC classification and quantification (CTC-Quant) algorithmic pipeline for our study. The pipeline is capable of high-throughput image data. It consists of 4 components. 1) A robust pre-processing module to remove image artifacts and standardize image intensities to ensure a fair comparison between images. 2) A segmentation and profiling module to extract individual cell segments and profile them for specific parameters including size, morphology and protein expression based on fluorescent labels. 3) A classification module using CTC features inspired by prior literature (8,18,19), enabling higher explainability of cell-type classification. It also offers more comprehensive classification of CTC subtypes, including epCTCs, bipCTCs and mCTCs as compared to other image classification solutions offered previously. It further differentiates between high-confidence CTCs versus background cells with similar staining patterns based on modelling the expression levels in healthy controls assumed to have minimal to no CTCs. 4) A cluster identification module to quantify clusters with three or more nuclei since clusters have been previously shown to raise metastatic potential. All components are described in detail in the Methods section of this paper.

## Results

### Short-time window analysis of CTC count enhance precision of treatment response prediction

CTC-Quant was applied towards the study of treatment response in lung cancer. Blood samples were taken from 11 unique lung cancer patients undergoing treatment after each cycle until they stopped responding. An increase in tumor size seen on a CT scan was defined as progressive disease (PD), while maintenance or decrease of tumor size was defined as stable disease (SD) and partial response (PR), respectively. Of the 11 patients, 5 were non-responding and 6 were responding at cycle 2. Of the 6 responders at cycle 2, only 3 continued to respond when the study ended (clinical data in Supplementary Table 1). All blood samples were prepared as per described (Methods section), imaged and analyzed using CTC-Quant (counts presented in Supplementary Table 2). To account for natural elevation of CTC counts in some patients, change in counts of readings (relative to baseline/alternative reference reading) were calculated and used for further analysis.

Slicing the longitudinal CTC count data into 2-cycle windows allowed for refined treatment response prediction. Window analysis was carried out to study CTC changes over an isolated period during the treatment course. Here, CTC count changes were calculated relative to the measurement taken 2 cycles before the cycle of interest. Each window’s change was associated with a response status (PD, PR, or SD) determined via CT scan, and batch-effect correction was performed to account for batch-effect arising from extracting ‘trendline sections’ from each patient. The grouping into high and low count groups was determined via the cutoff that best maximizes the difference between PD and PR responders. Changes in CTC counts between PD, PR and SD response windows for all CTC subtypes were found to be statistically significant (p<0.05) as presented in Figure 1A. While noting that the absolute threshold may not generalize beyond our preliminary analysis, such evidence does motivate further exploration of the use of CTC to chart treatment response.

**Fig. 1.**
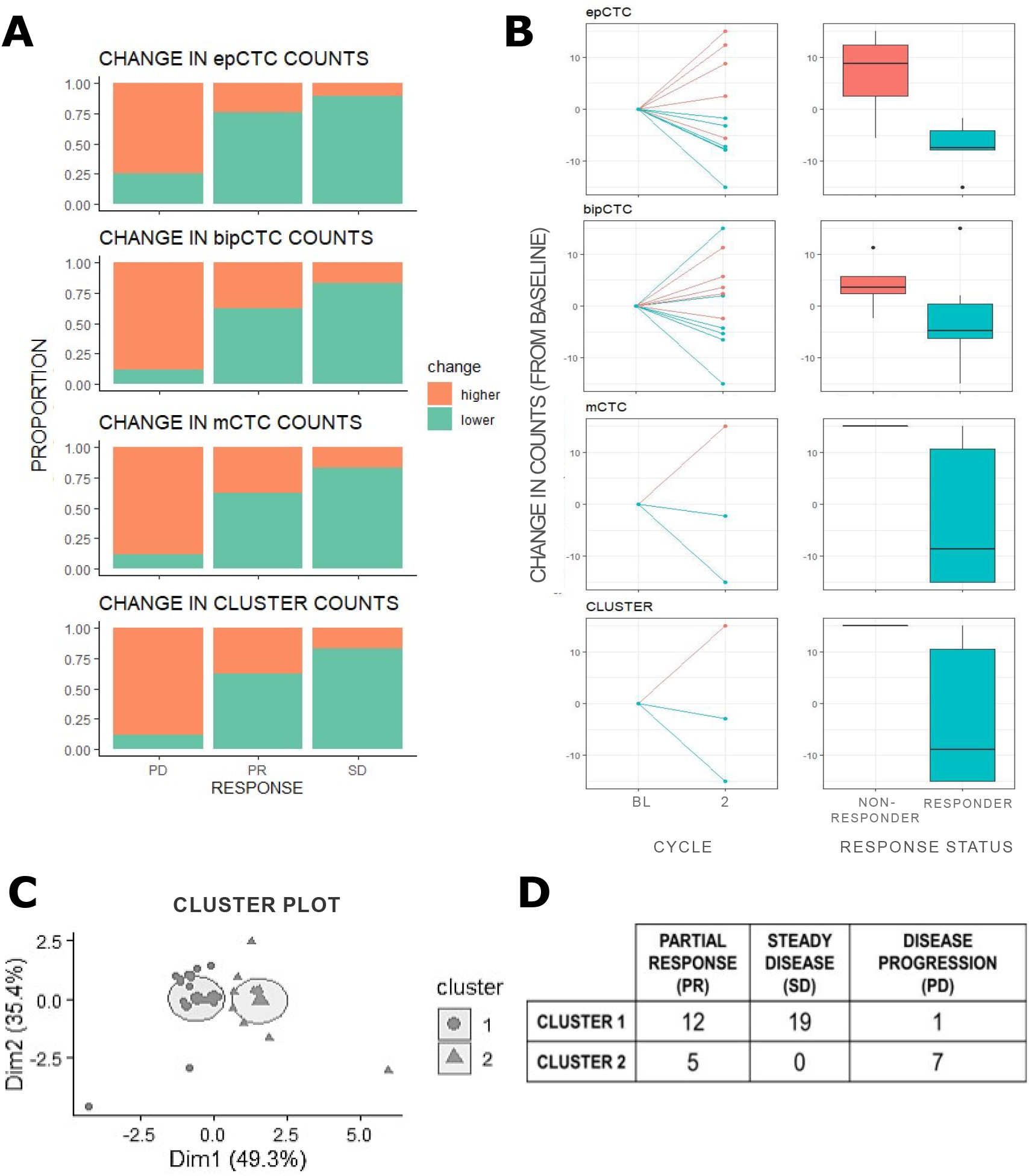
Change in CTC subtype counts is associated with disease response. Three subtypes of CTCs were quantified: epithelial (epCTC), biphenotypic (bipCTC) and mesenchymal (mCTC). mCTC clusters identified as cell clusters with >2 cells within a close periphery were also quantified. **(A)** In our short-time window analysis, CTC count changes are calculated relative to measurements taken two chemotherapy treatment cycles before. Each window is associated with either of Progressive Disease (PD), Partial Response (PR), and Steady Disease (SD) determined via a CT scan. A larger proportion of PD windows were found to have higher numbers of CTCs, while a larger proportion of PR and SD windows had lower numbers of CTCs. **(B)** Starting from baseline, patients that responded to chemotherapy (PR and SD patients) after two cycles of treatment and non-responders (PD patients) were found to be differentiated by their CTC count changes (relative to baseline). Responders experienced a decrease (or no change) in all CTC subtypes whereas non-responders see an increase in all CTC subtypes. Figures were clipped at (−15,15) for visualization purposes. **(C)** Unsupervised clustering using Kmeans on epCTC, bipCTC, mCTC and cluster count found two main clusters. **(D)** Majority of PR and SD windows were part of Cluster 1 while majority of PD windows were part of Cluster 2, confirming differentiable trends in CTC subtype counts in responders vs. non-responders.

In a comparison of responders against non-responders at treatment cycle 2, our analysis reveals a trend in which treatment responders see a decrease (or no change) in all CTC subtypes, whereas non-responders see an increase in all CTC subtypes (Figure 1B). Quantifying this trend, we find that at least 67% of the responders reflected a decrease in epCTC, bipCTC, mCTC and cluster counts, as compared to at most 20% of non-responders (Supplementary Table 3).

Unsupervised clustering by Kmeans was carried out on the change in counts (2-cycle window), and found to be best grouped into 2 clusters (Figure 1C). We find that 71% of PR and 100% of SD windows were part of Cluster 1, while 87.5% of PD windows were part of Cluster 2 (Figure 1D), indicating a differentiable trend between responders and non-responders in CTC count change (2-cycle window).

Window analysis of short and precise periods of longitudinal CTC count data allows for a more curated, refined and insightful understanding of patients’ near real-time response (Figure 2). Patients who were still responding at the end of their second cycle continued to be followed until they stopped responding or the study ended. In Figure 2, patient P01 who responded to all treatment cycles (including the last measurement taken) was found to present both a decreasing and saw-toothed response, despite having a sustained stable disease that was not indicative of response to treatment. A computation of change in counts relative to baseline was similarly found to be limited in providing observable correlation with treatment response. This was especially so as treatment time progressed and the period between the latest measurements and baseline measurements increased. In contrast, the 2-cycle window method of computing CTC count changes appeared to be able to capture a more up-to-date response, presented by a fairly consistent measurements throughout P01’s stable disease.

**Fig. 2.**
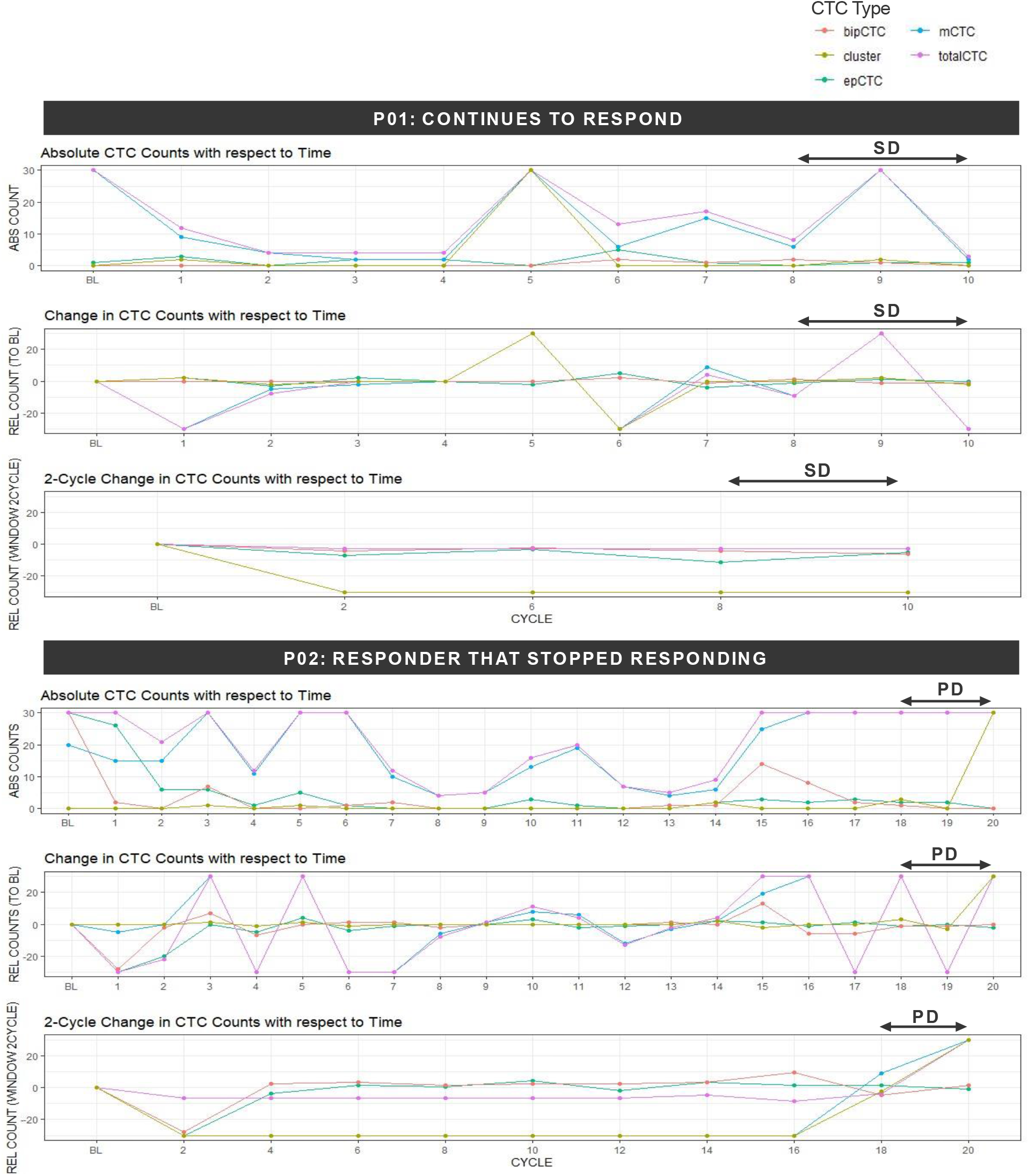
Comparison of longitudinal absolute counts, relative (to baseline) counts and window analysis. Patients undergoing chemotherapy were followed over time and had biopsies taken after every treatment cycle until they stopped responding (i.e. experienced progressive disease (PD)). The longitudinal chart of absolute counts with respect to time fluctuates drastically and does not present observable correlation with treatment response. Change in counts (relative to baseline) similarly is limited in capturing the dynamic nature of CTC count changes and is limited in providing insight to response. In contrast, 2-cycle window quantification of CTC count changes reveals a clearer distinction between the decrease/stability during response, and an increase when patients who were previously responding experienced progressive disease.

P02 was an initial responder that experienced PD and had stopped responding at the last measurement. When monitoring P02’s CTCs, a similar challenge was faced when studying absolute and change (relative to baseline) data over time. Once again, longitudinal patterns of CTC counts did not have observable signs of treatment response or non-response. Instead, sectioning out short periods of treatment times clearly visualized a spike in CTCs when P02 experienced progressive disease. Compared to a time-series chart of raw absolute CTC counts and an even change in counts (relative to baseline), the window analysis applied captures the dynamic nature of CTC count fluctuation with respect to treatment response from cycle to cycle. In this way, CTC count changes are contextualized to a patient’s response to the on-going therapy they are concurrently undergoing. This localization of measurements to treatment period allows for refined real-time prognostication of treatment response further improves the effectiveness of CTC monitoring in disease management.

The trend of responders reflecting a decrease in CTC counts and non-responders reflecting an increase (change>0) in CTC counts relative to baseline is also seen in an analysis of 30 nasopharyngeal cancer patient samples (with complete post-cycle measurements and response data). 52%, 52% and 60% of responders showed no-change/decreased (change<0) epCTC, bipCTC and mCTC counts, respectively. 60%, 80% and 60% of non-responders instead showed an increase in epCTC, bipCTC and mCTC counts, respectively (Supplementary Table 3).

### bipCTC and mCTC counts are associated with tumor staging

Blood samples were taken from 41 breast cancer patients. Of the 41 breast cancer samples, 26, 11 and 4 patients were found to have stage IV, III and II cancers, respectively (clinical data in Supplementary Table 1). A larger proportion of advanced cancer patient samples were found to have high bipCTC and mCTC counts (p<0.05). Positive rates of bipCTCs and mCTCs were highest in stage IV breast cancer patient samples at 42% and 85%, respectively. In comparison, positive bipCTC and mCTC rates were lower in stage II/III breast cancer patient samples at 0– 25% and 25–55%, respectively, as presented in Table 1. This is an expected observation since bipCTCs and mCTCs are subpopulations that have demonstrated mesenchymal-epithelial-transition (EMT) identified as a critical process for cancer progression. Previously established correlation between these two EMT subtypes and distant metastasis (16) hence supports the trend that later-stage breast cancer patients who experience metastasis then present more bipCTCs and mCTCs. In addition, 31% of stage IV breast cancer samples were found to have at least one mCTC cluster, as compared to 9% of stage III samples and 0% of stage II samples. Since CTC clustering is an adaptive mechanism to enhance survival during transport that promotes metastatic efficiency by 20–100 times (17,20), it is understandable that patients with more aggressive cancers are more likely to present mCTC clusters.

**Table 1.** CTC subtypes positive rates by tumour staging in breast and nasopharyngeal cancer. Breast cancer patients were found to express higher numbers of mesenchymal-like CTCs and clusters in later stages of cancer, while nasopharyngeal cancer patients were found to express higher numbers of the same CTC subtypes and clusters in earlier stages of cancer.

| BREAST CANCER STAGING |  |  |  |
| --- | --- | --- | --- |
| CELLTYPES | STAGE 4 (n=26)<br>PROPORTION HIGHER | STAGE 3 (n=11)<br>PROPORTION HIGHER | STAGE 2 (n=4)<br>PROPORTION HIGHER |
| epCTC | 0.12 | 0.18 | 0.25 |
| bipCTC | 0.42 | 0.00 | 0.25 |
| mCTC | 0.85 | 0.55 | 0.25 |
| Total CTC | 0.81 | 0.64 | 0.25 |
| clusters | 0.31 | 0.09 | 0.00 |
| NASOPHARYNGEAL CANCER STAGING |  |  |  |
| CELLTYPES | STAGE 4 (n=9)<br>PROPORTION HIGHER | STAGE 2,3 (n=22)<br>PROPORTION HIGHER | STAGE 1 (n=7)<br>PROPORTION HIGHER |
| epCTC | 0.00 | 0.09 | 0.29 |
| bipCTC | 0.22 | 0.41 | 0.00 |
| mCTC | 0.78 | 0.59 | 1.00 |
| Total CTC | 0.78 | 0.91 | 1.00 |
| clusters | 0.11 | 0.18 | 0.29 |

A similar analysis was run on 38 of 41 nasopharyngeal cancer patients with complete tumor staging records but was found to exhibit a different abundance of CTC subtypes at each cancer stage. Of the 38 patients, 6, 22 and 7 patients were diagnosed with stage IV, II/III and I cancer, respectively. Stage I patient samples were found to have the largest proportion of high mCTC counts and clusters than stage II/III or IV. Interestingly, a higher proportion of early-stage nasopharyngeal cancer patients have high mCTC and cluster numbers (agreeing with a similar trend previously reported in (21)) directly contrasting breast cancer in which late-stage patients have more mCTCs and clusters. Furthermore, stage II/III patient samples were found to have the highest proportion of high bipCTC counts at 41%, as compared to 22% and 0% in stage IV and stage I patient samples, respectively. Unlike breast cancer whereby stages with high mCTCs also had high bipCTCs, nasopharyngeal cancer had a distinct pattern where stages with high mCTCs had lower bipCTCs, and vice versa.

bipCTCs and epCTCs are correlated with EBV DNA levels that is a prognostic factor for tumor staging in nasopharyngeal cancer. The patients had their EBV DNA levels measured at two time points. The first EBV DNA measurements for each patient were binned into three groups - <2200, >2200, and undetectable. Effect sizes were calculated between CTC subtype counts and EBV groups. bipCTCs and mCTCs were found to have large effect sizes with etasq 0.14 and 0.13 respectively, and a cohens f of 0.40 for both. CTC clusters were also found to have a medium effect size of etasq 0.07 and cohens f of 0.27.

Tumor staging today estimates the extent of disease through imaging tests such as CT scans and X-rays, or pathological tests using biopsies. The ability to use CTCs as a simple minimally invasive test to give a signal of cancer staging (albeit largely indicative rather than deterministic due to the heterogenous nature of cancer) eliminates the need for patient exposure to ionizing radiation or invasive surgical procedures. Mesenchymal-like CTC subtypes in particular may help to meaningfully identify patients at risk for further screening.

### CTC-Quant achieved excellent agreement with expert opinion

Our classification pipeline was found to achieve high levels of agreement with expert-generated labels. When identifying epCTC or background epithelial cells, bipCTC or other background cells and mCTC cells (see CTC-Quant counts in Supplementary Table 2), CTC-Quant labels achieved a high agreement (minimum average Kappa score of 0.85) with the two experts (Table 2). Since the experts’ labels had a level of disagreement between themselves (Supplementary Table 4), only cell segments that were labelled similarly by both experts were used to calculate specificity, sensitivity, false positive rates (FPR) and false negative rates (FNR) for the following expert and CTC-Quant disagreement analysis. CTC-Quant achieved extremely high specificity of minimally 0.98 across all cancer types, but sensitivity ranged between 0.66 and 0.99 (wide sensitivity range to be explored below).

**Table 2.** CTC-CQ validation against expert labels for four data sets – breast, nasopharyngeal and lung cancers, and healthy controls.

|  | METRICS | BREAST<br>CANCER<br>(n = 2072) | NASOPHARYNGEAL<br>CANCER<br>(n = 2372) | LUNG<br>CANCER<br>(n = 2288) | CONTROL<br>(n = 2052) |
| --- | --- | --- | --- | --- | --- |
|  | Average<br>Cohen's Kappa | 0.89 | 0.88 | 0.84 | 0.90 |
| epCTC /<br>background<br>Epithelial | Specificity | 1.00 | 0.99 | 0.99 | 0.99 |
|  | Sensitivity | 1.00 | 1.00 | 0.98 | 1.00 |
| bipCTC /<br>other<br>Background | Specificity | 1.00 | 1.00 | 1.00 | 1.00 |
|  | Sensitivity | 0.83 | 0.89 | 0.93 | 0.94 |
| mCTC | Specificity | 1.00 | 0.99 | 0.98 | 0.98 |
|  | Sensitivity | 0.66 | 0.92 | 0.83 | 0.98 |

Further analysis of label differences between CTC-Quant and the experts reveals that CTC-Quant is more deterministic. Segments that were labelled CTC by experts but not CTC-Quant were found to not fit the literature definitions of the relevant CTC subtypes (presented in the Methods section). Differences in labels were mainly due to different biases on vimentin (VIM) staining and cell diameters. When profiled (Supplementary Table 6), we find that most often experts would label the VIM staining differently when the staining intensities lie close to the cutoff threshold (value obtained as described in Methods). For example, epCTCs were often labelled as bipCTCs or mCTCs during manual labelling, despite having lower, non-significant average vimentin expressions or when their cell diameters were smaller than that reported in literature (presented in Methods section). This indicates that experts may be more likely to identify false-positive CTCs. In contrast, CTC-Quant’s expression cutoff (based on a large population of cells) and size filtering allow for deterministic classification and suggest that the actual sensitivity is higher than when calculated against expert labels.

Going beyond manual labelling-type identification of CTC subgroups, CTC-Quant achieves higher specificity by separating high-confidence CTCs from background cells. Based on the assumption that healthy control samples should have minimal to no CTCs, we model the cytokeratin (CK) expression of cells from the healthy control samples as ‘background expression’ and determine average CK expressions above the threshold to be significant CK signals and high-confidence CTCs (elaborated in Methods section). This means that the final CTC counts of CTC-Quant (quantifying high-confidence CTCs) will be lower than manual counts (quantifying all cells with the relevant staining patterns regardless of expression levels), and may help to mitigate the low purity and higher false-positive rates faced during the use of the microsieve and manual labelling.

## Discussion

The adoption of CTCs as a prognostic biomarker is alluring as it creates opportunity for frequent minimally invasive and continuous monitoring of treatment response and prognostication. As presented earlier, our analysis reveals that CTC count changes after treatment differentiate responsiveness and non-responsiveness towards the treatment received. Concurrent analysis of CTC count changes with each treatment cycle further hints at the potential ability to carry out near real-time prognostication of treatment response. In addition, absolute CTC counts, particularly those with mesenchymal characteristics, are found to be strongly associated with tumor staging, which can aid in risk stratification to triage patients for pre-emptive scans of early-stage clinical assessment.

The current day understanding of CTC counts associations with response and survival still requires refining. Time series study of CTC changes over time has been made possible with the advent of a wide array of CTC enumeration and analysis techniques. Going beyond the aforementioned associations found with progression-free and overall survival rates, charting CTC counts over treatment cycles has revealed various types of possible response patterns (22) and observable trends (23) that indicate disease progression versus response of patients undergoing therapy. These interesting findings have proven the utility of CTCs in clinics for the general understanding of a patient’s potential survival time and understanding response retrospectively. However, there is still limited ability to carry out real-time determination of treatment response. Achieving real-time understanding of each patient’s response to therapy is essential for the practical implementation of CTC monitoring in clinics. Real-time insights of response status can empower physicians to quickly evaluate and tailor the most suitable treatment options for each patient moving forward to greatly reduce unnecessary toxicity from unsuitable treatments. Given this, a refined understanding of CTC changes capable of providing close to real-time information on response is highly anticipated.

In this respect, studying small, localized short-time windows of longitudinal CTC count trendlines provides refined and useful insights, enhancing personalized near real-time disease monitoring. Two key strengths of this analysis include the quantification of scaled CTC count changes (relative to previous readings), and small window analysis that provides an indication of treatment response during that specific time period. The former helps to account for bias in natural elevation of CTCs in some patients relative to others with similar diagnosis, and aids generalizability when studying CTC changes with respect to response in a population. The latter sections short periods of interest (2-cycle period in this analysis) and studies patient’s CTC change given their response every 2 treatment cycles. This window analysis enables us to study response and the associated CTC count change during short, isolated periods of therapy. Given that patients do not always experience consistent SD or PR, extrapolation of these insights is extremely beneficial in allowing clinicians to obtain refined and close to real-time indications for when to tailor treatment types, frequency and dosages to better improve patient outcomes. Extending the effort of previous work, this finding takes a step forward toward achieving the personalized and real-time treatment monitoring and is worth further investigation.

Our small sample size used for the window analysis is a notable constraint limiting the generalizability of our findings. Where a larger cohort of patients is available for study, it would be interesting to evaluate the generalizability of our observed trends to the larger population of lung cancer patients undergoing treatment. In addition, only our lung cancer patients had complete time-series CTC measurements and relevant CTC scans available, limiting the breadth of cancer types to which we could apply this window analysis. Given that different cancers are expected to have different CTC presentation patterns and abundances, it is possible that the computation of CTC changes over small timeframes may have varying levels of usefulness in aiding the evaluation of treatment response in different cancer types. A broader study over a larger sample of patients and across different cancers would be advantageous in better determining the generalizability and applicability of this finding. Such an extension would be particularly desirable for better modeling of short-period treatment response to predict and recommend best treatment options for patients of which we have yet to achieve in this study.

The duration of extracted windows for which CTC count changes were computed for analysis is also worthy of further consideration. Sampling frequency has been raised as a point of contention (22) due to the large variability in CTC numbers over time. This means that the duration between each sample or the duration at which changes in CTC counts are computed over may have a significant impact on the ability to fully capture meaningful changes in CTC counts. Moving forward, a refined analysis to determine an optimal sampling frequency and duration of analysis windows is imperative in striking a balance between an unfortunate omission of meaningful trends and an undesirable inclusion of fluctuation noise.

Absolute CTC counts also have utility in tumor stage determination. In this study, higher proportions of late-stage breast cancer patients were found to present higher numbers of mesenchymal-like CTC subtypes and CTC clusters than early-stage patients. The asymmetry between CTC presentation patterns at different stages in nasopharyngeal cancer and breast cancer hints at the idea that the two main models of metastasis (i.e. the linear and parallel progression models) may apply in different cancer types. In the linear model, cancer cells undergo a series of mutations and selections for the fittest (24,25) within the primary tumor before cancer cells leave the tumor and seed secondary growths. Based on this idea, CTCs (especially bipCTCs and mCTCs) are expected to be observed in later-stages of cancer after the cells have undergone successive rounds of proliferation within the primary tumor. This trend was observed in the analysis of breast cancer patients earlier, suggesting that the linear progression model may be applied in the explanation of breast cancer metastasis. Indeed, other studies of breast cancer patients have also supported the linear progression model from advanced clonal subpopulations (26,27,28). In contrast, the parallel progression model postulates that cancer cells have metastatic potential early on in disease development (25,29). Hence tumor clones can start to seed and metastasize at secondary sites simultaneously with primary lesion growth. Given this, detecting higher proportions of early-stage nasopharyngeal cancer patients presenting more mesenchymal-type CTCs that are highly metastatic could indicate that parallel progression (where metastasis has been suggested to begin much earlier) occurs in the development of nasopharyngeal cancer.

These results also promote the idea of utilizing CTC tracking to identify development of early-stage into late-stage cancer. This can be used in tandem with treatment response to understand when patients respond poorly to treatments and are experiencing disease progression and developing later-stage cancers. However, differences in mesenchymal-like CTCs across stages in nasopharyngeal cancer compared to breast cancer highlight the heterogeneity of CTCs across different cancer types. This suggests that the developmental mechanisms of early-to late-stage cancer in nasopharyngeal and breast cancers could be very different and highly complicated. As such, further experiments (such a multi-omic analysis of samples over time at different stages of cancer progression) are required to better elucidate the potential mechanisms of interest and may also help in differentiating metastatic development across different cancers.

The massive uptick in the amount of image data collected holds great potential to unlock extensive supplementary insights towards the clinical application of CTCs. Yet, the current reliance on trained operators to manually screen the large number of images limits the feasibility of efficient and effective high-throughput analysis. The manual classification and quantification is tedious, and suffers from high inter-operator variability (30,31). The subjective nature of classification by eye further makes identification and classification of CTC subtypes challenging.

Many solutions have been proposed to address complexities arising from manual CTC classification and quantification in images. Software-based solutions can be grouped into machine learning image analysis and heuristic-based classification (summarized in Supplementary Table 7). Examples of machine learning image analysis and cell classification solutions include ACCEPT (31,32) and other Convolutional Neural Networks (CNN) (33,34,35). Heuristic-based classification conducts classification based on biophysical properties of cell segments (36). Several groups have gone even further to develop whole imaging systems (37,38,39) for the study of CTCs, including the single FDA-approved CellSearch system (40). Despite the reported high levels of accuracy achieved by these systems, the inertia against wide-spread adoption of available solutions is possibly due to low explainability, ability to detect only specific CTC subtypes or even non-accessibility to use them. Models that can explain their predictions are preferred to ensure that classification rules are biologically relevant and trustworthy. Some machine learning models, especially the commonly used black-box CNN-based classifiers are difficult to explain, and an alternative model may be preferred by scientists. Furthermore, several subtypes of CTCs were reported to have varying levels of effect on metastasis. However, many of these image analysis software and classifiers were trained to identify only a specific subset (usually EpCAM+/CK+), limiting the ability to study these CTC subsets of interest. Moreover, solutions that require users to acquire a specific imaging tool or an imaging system may be a barrier-to-adoption. This is especially so when some of the imaging systems have yet to be made readily available on the market, and code-based analysis pipelines are not made readily available for replication.

An image analysis solution capable of reducing human-intervention complexities that is explainable, capable of elucidating CTC subtypes for more specific studies, and made available for use would better enable others interested in studying CTCs to use the solution. CTC-Quant meets these conditions and is particularly useful in addressing prior concerns regarding the adoption of various image analysis methods for CTC studies. We note that universally true ground-truth labels have yet to be generated for comparison across various methods, and manual labels (despite their inter-observer variability) have been taken to be the traditional ‘ground-truths’. While CTC-Quant’s classification heuristics were generated from extensive findings in literature, its accuracy at classification was also validated against subjective manual labels. The use of known pure CTC subtype images (though challenging to obtain) may help with further validation and potentially boost CTC-Quant’s reliability.

We take caution in the application of our classification algorithm toward a larger population of patients. Given that cancer is a very heterogenous disease, further refining to preserve the robustness of our algorithm when scaling up is foreseen. More comprehensive normalization can be implemented to ensure fairer comparison of different sample images, especially if they are acquired by alternative machines. In profiling, an inclusion of alternative indexes such as sphericity, roundness or roughness can be used to capture cell morphology more precisely for classification. For further contextual information, distance metrics between each cell and the cells in their periphery can be computed so as to provide more information of possible cell interactions and clustering. Validating the classification and quantification method on a larger sample group would certainly help to generalize the classification heuristics toward the disease population.

Routine yet minimally invasive liquid biopsies from patients makes CTC tracking a promising strategy for monitoring treatment efficacy and resistance. Strengthened by reliable and accurate methods to analyze CTC images for high-throughput analysis, further efforts to study precise periods of treatment response and early diagnosis using CTCs could greatly reduce unnecessary toxicity from unsuitable treatments, increase the matching of patients to effective and personalized treatments, and improve overall patient outcomes.

## Materials and Methods

### Data

All individuals provided written informed consent for blood-taking. Patients with lung, breast and nasopharyngeal cancers were recruited from Changi General Hospital, National University Hospital and National Cancer Centre between 2013 to 2018. 5 independent CTC image datasets were utilized for this study – 3 lung cancer patient cohorts consisting of 40 (41), 14 and 11 patients; 1 breast cancer patient cohort consisting of 41 patients; 1 nasopharyngeal cancer patient cohort consisting of 41 patients (42); and 1 group of 19 healthy controls.

The 40-sample lung cancer dataset (41) was used in the training of the CTC-Quant algorithm which was later tested on the second unseen 14-sample lung cancer dataset. The breast cancer and nasopharyngeal cancer datasets were split 70:30 into training and testing datasets for the generation of our classification and quantification method. Microfabricated silicon microsieves with round pores (9-10 μm in size) were used for CTC isolation. These microsieves have been previously described and validated for highly efficient tumor cell capture (42-45). Briefly, 1 mL of whole blood was diluted with 2 mL of PBS/EDTA/BSA buffer solution, and then passed through the sieve at 0.5 mL/min using a peristaltic pump. Cells were fixed with 4% paraformaldehyde, then permeabilized with 0.1% triton-X before incubation in the antibody mixture containing DAPI, CD45-Alexa Fluor 647 (1:25; Bio-Rad MCA87A647), Pan-Keratin-Alexa Fluor 488 (1:100; Cell Signaling Technologies 4523S) and Vimentin-TRITC (1:100; Santa Cruz Biotechnology sc-6260) for 1 hour at room temperature. Three washes of 1 mL of PBS/EDTA/BSA were used between each step. The sieve was then scanned and imaged automatically using a fluorescent microscope at 20×.

### Modules 1 and 2: Image pre-processing, segmentation and profiling

Upstream image pre-processing followed by segmentation and profiling was achieved with the first two modules in our analysis pipeline (Figure 3A).

**Fig. 3.**
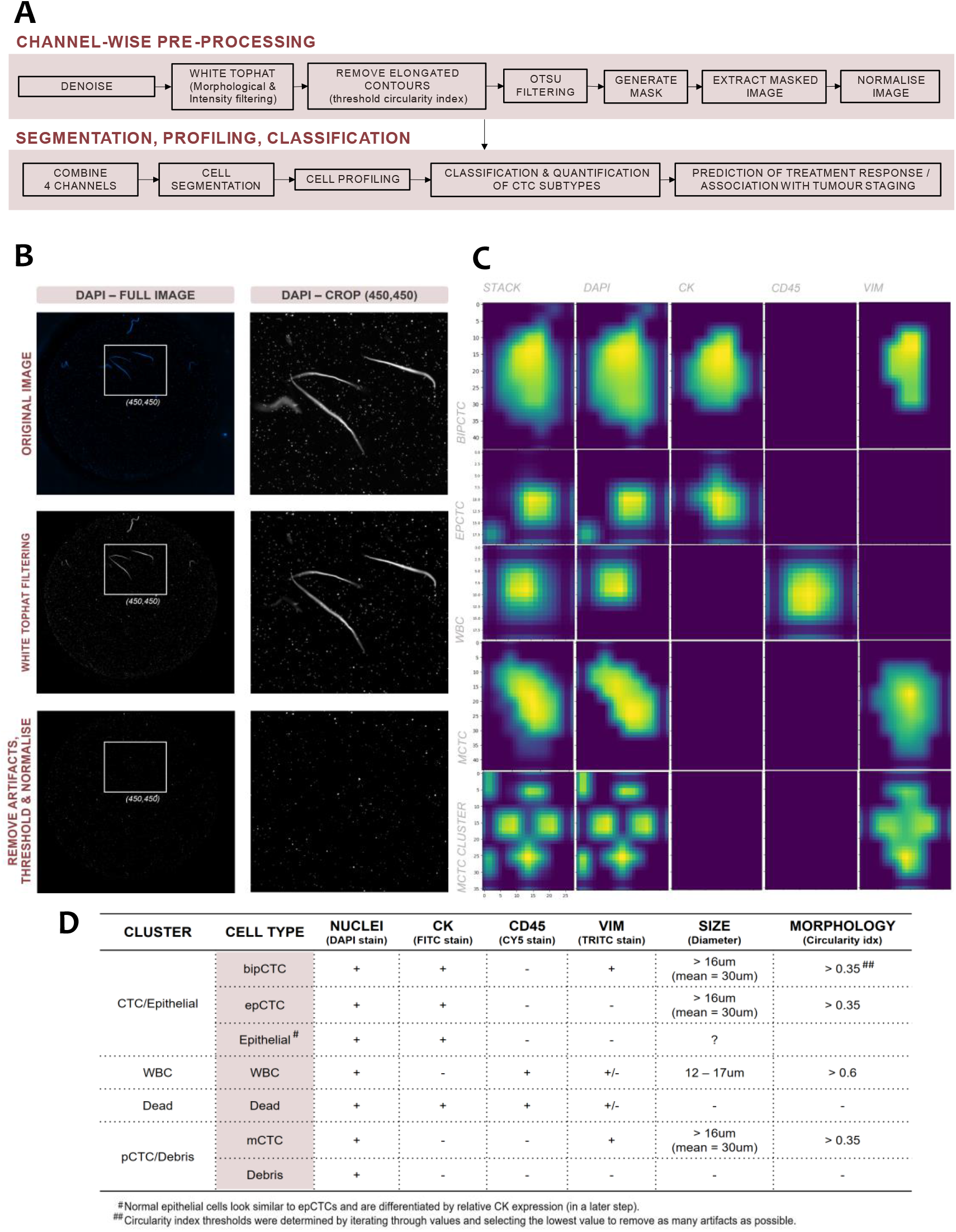
CTC-Quant classification pipeline, visualization of image processing steps, cell segment visualization and classification heuristics. **(A)** Flowchart demonstrating the steps taken to classify and quantify CTC subtypes from microscope images starting from image pre-processing to segmentation to cell profiling to classification and quantification. Following which, the algorithm was used for the analysis of chemotherapy treatment response analysis and tumor staging correlation. **(B)** A visualization of the image pre-processing steps clearly presents how the image pre-processing steps are essential toward removing sample preparation and imaging artifacts, as well as extracting meaningful signals. **(C)** Following pre-processing and segmentation, individual cells are segmented and profiled for their cell area, cell diameter, circularity index and average protein expression (signals) in each channel. **(D)** The segmented cells are then classified according to heuristics determined from literature including staining intensity and expected cell sizes. Cell morphology is also used as a filter to remove artifacts that are much more elongated than the expected cells.

Module 1: Pre-processing was performed channel-wise (Figure 3B). The raw images were denoised via total variation regularization. Following that, they were filtered using the white-top hat transform to remove large bright spots due to acquisition defects. Elongated preparation artifacts were further removed through thresholding circularity index of contours detected. Otsu thresholding was then used to mask and extract meaningful signal. Adaptive histogram equalization was finally applied to normalize the intensity for all images that had undergone pre-processing.

Module 2: All four channels were stacked and flattened before the watershed segmentation algorithm was used to extract potential cell segments (Figure 3C). To account for touching cells, a minimum distance of 2 × minimum cell radius (9 μm) between nuclei (DAPI channel) staining was set. Each potential cell segment was then profiled to obtain biophysical measurements including cell area, cell diameter (calculated from cell area assuming cells were circular), circularity index, average nuclei expression, average CK expression, average CD45 expression and average VIM expression. These biophysical measurements were essential for cell type classification (Figure 3D).

### Module 3A: Identifying 4 main cell clusters

All potential cell segments without DAPI signals and with overall diameters of < 9 μm were filtered out of the classification pipeline as all cells of interest were expected to have positive nuclei staining and to be larger than a white blood cell (WBC). By conventional definition, a CTC is larger than a WBC, stains positively for epithelial markers such as EPCAM and/or CK, and stains negatively for WBC markers such as CD45 (8). Hence, remaining cell segments were first clustered via Kmeans clustering into 4 main clusters based on the presence of CK and CD45 staining (Figure 4A). Cell segments that were CK-CD45+ were clustered as WBCs; CK+CD45-as epCTC/bipCTC/normal epithelial cells; CK+CD45+ as dead cells; and CK-CD45-as mCTC/debris (Figure 4A).

**Fig. 4.**
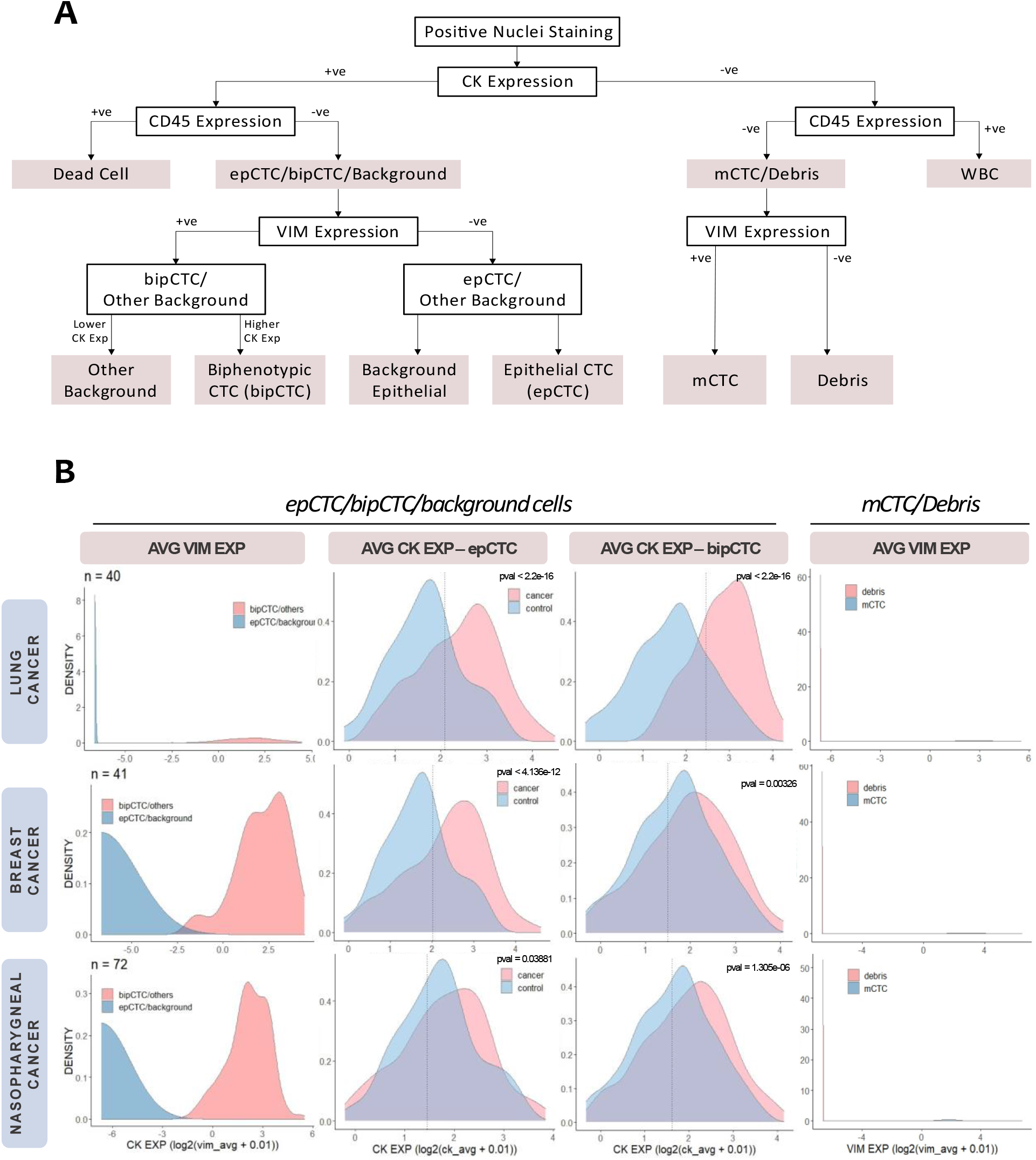
Classification decision tree and density plots enabling differentiation of CTC subtypes. **(A)** Flowchart of CTC subtype classification. **(B)** Density plots of average VIM expressions in lung, breast and nasopharynfeal cancers reveal clear VIM+ and VIM-clusters. CK expressions for epCTC/bipCTC/background cells and mCTC/debris in the cancers are also found to have statistically significant differences from cells in healthy controls. Since healthy controls are presumed to have minimal (or even no) CTCs, these distributions can help to further extract meaningful signal of cancer cells from background cells. Cutoffs to best differentiate cells with meaningful signals from background cells are obtained via general mixture modeling (GMM).

### Module 3B: Differentiating epCTC and bipCTC and from background cells

Recognizing the fact that CK proteins are markers of normal epithelial differentiation and not specific to CTCs, we looked further into differentiating CTC subtypes from background cells.

There are two epithelial-type CTC subtypes. The first subtype is the epCTC that stain positively for DAPI and CK only. The second subtype is the bipCTC that stain positively for DAPI, CK and VIM. bipCTCs refer to CTCs from epithelial origin that have undergone a certain level of epithelial-mesenchymal transition (EMT), resulting in their expression of Vimentin (a canonical biomarker of EMT). Given this, the CK+CD45-cluster is further subdivided into VIM+ or VIM-sub-clusters through Kmeans clustering (see decision-making flowchart in Figure 4A, and clustering results based on average VIM expression in Figure 4B).

We compared the average CK expressions of CK+CD45-Vim-(epithelial-like) cells from cancer samples (n = 40 lung cancer samples; n = 41 breast cancer samples; n = 72 nasopharyngeal cancer samples) to healthy control samples (n = 19). Based on the reasonable assumption that samples from healthy patients should have fewer CTCs (even close to none), we found that cells from the cancer samples have a statistically significantly higher average expression (than cells from the healthy controls) across all our cancer data sets (non-parametric t-test pvals < 0.05, Figure 3B). Gaussian mixture models were used to group cell segments into high-CK and low-CK expressing groups for further CTC subtype vs. background cell classification. The former group was also deemed the high-confidence CTC group as the average CK expression was of higher confidence to be a significant signal.

Previous studies have also found that CTCs generally have a larger cell diameter than healthy cells, and are minimally 16 μm in diameter. Hence, CK+CD45-VIM-cells with higher average CK expressions and diameters ≥ 16 μm (18,19) were classified as epCTCs, while all others were classified as background epithelial cells. Likewise, CK+CD45-VIM+ (biphenotypic) cells were also found to have a statistically significantly higher average CK expression when compared against those from the healthy control samples (non-parametric t-test pvals < 0.05, Figure 3B). CK+CD45-VIM+ cells with higher average CK expressions and diameters ≥ 16 μm were classified as bipCTCs, while the others were deemed to be background.

### Module 3C: Differentiating mCTCs from debris

mCTCs are a subset of CTCs considered to have undergone EMT and have lost their expression of CK proteins. Hence, this subset of CTCs is differentiated from debris based on the presence of vimentin (Figure 4A). CK-CD45-cells were subdivided into VIM+ or VIM-subtypes based on their vimentin staining (Figure 4B). Lastly, CK-CD45-VIM+ cells with cell diameters ≥ 16 μm (18,19) were classified as mCTCs, while all other CK-CD45-cells were classified as debris.

### Module 4: Identifying mCTC clusters from single mCTCs

mCTC clusters have been shown to cause aggressive metastasis. Hence, it is of great interest to differentiate these clusters from single mCTCs. mCTC segments with 3 or more segmented nuclei stains within a close radius were labelled and counted as a cluster (Figure 3C).

### CTC-Quant Validation

Approximately 2000 cell segments from each testing data set (including the control, lung cancer, breast cancer and nasopharyngeal cancer) were separately labelled by two trained experts. Since CTCs are rare events in blood samples, all cell segments with CK expression were included for validation by the experts, while non-CK-expressing cell segments were randomly selected to be included for expert labelling. The expert labels were first compared using a confusion matrix, and their interrater reliability was evaluated using Cohen’s Kappa. The expert labels were then compared against the CQ-CTC generated cell type classification labels. epCTCs and bipCTCs were classified together with their healthy counterparts as it was difficult to discriminate them based on their staining patterns by eye.

### Statistical Analysis

Distributions of expression values were checked for normality using the Shapiro’s Test for Normality. Where the assumptions for a normal distribution were not met, the Kruskal-Wallis test was used to calculate pvalues for comparison. CTC counts were studied, and outliers were treated with one-sided windsorization (to the 90th percentile). Change in CTC counts was calculated to account for inherent inflation of CTC numbers in patients by subtracting the CTC counts at a reference time point from the counts at the time point of interest. Batch-effect correction was carried out using ComBat from the ‘sva’ library in R (46,47).

When comparing clinical groups such as tumor staging or treatment responders vs. non-responders, samples are classified into ‘higher CTC count’ and ‘lower CTC count’ groups and the Fishers Exact test was used for statistical comparison. Thresholds for this classification by absolute counts were obtained by iterating through all possible cutoff values and selecting the one that maximizes the difference between groups.

Unsupervised clustering of localized 2-cycle windows was done using Kmeans. The elbow method was used to determine the optimal number of clusters, which was two in this case.

## Data Availability

All data are available in the main text or supplementary material.

## Author contributions

Conceptualization: CML, KL, JM Methodology: CML, KL, JM

Materials: TMC, AT YSY, WN, DC, SWW

Clinical Sample Processing: JS, JV, HM

Expert Validation: MN, NBMM

Data Analysis & Visualization: CML

Supervision: KL, JM, JYY

Writing — original draft: CML

Writing — review & editing: CML, KL, JM, JYY

## Data and materials availability

All data are available in the main text or the supplementary materials. The CTC-Quant code has been made open-source on Github. Simulated image data is also available.

## Declarations

This study was approved by the SingHealth Centralised Institutional Review Board for Changi General Hospital and National Cancer Centre and the NHG Domain Specific Review Board for National University Hospital.

